# Is population anxiety associated with COVID-19 related hospitalizations and deaths? A study protocol

**DOI:** 10.1101/2020.07.16.20155457

**Authors:** Frederik Feys, Abdallah Naser

## Abstract

**BACKGROUND:** The nocebo effect is any harmful reaction following a negative suggestion. Anxiety can be seen as a manifestation of a nocebo context. The psychological stress of the COVID-19 pandemic led some people to experience COVID-19 symptoms, which were not actually related to a COVID-19 infection. A fundamental goal during the COVID-19 pandemic is to limit the COVID-19 related demand on healthcare systems and to minimize COVID-19 related deaths. This exploratory study aims to determine to what extent the anxiety in the population is related to the number of covid-19 related hospitalizations and deaths.

**METHODS:** We will quantify the magnitude of the relationship between population anxiety and hospitalizations / deaths. Anxiety will be assessed using the results of the most frequently used anxiety measuring scale. Official websites of governments will be screened to determine hospitalizations and deaths.

Studies will be included if they had at least 100 respondents, used a validated anxiety scale, reported on the general population of a country, and were conducted during the Covid-19 pandemic. A search strategy will take into account the limited resources for this study and will be used to search Pubmed, MedRXiv and PsychRXiv. Screening will take place at two levels: abstracts and titles, followed by full text reports. One researcher will extract data which will be double checked by a second researcher.

We will perform weighted OLS regression to quantify the relationship of anxiety and covid-19 related hospitalizations / deaths.

**DISCUSSION:** Covid-19 related measures can carry a significant social cost and risk of unwanted effects; it is therefore important to assess the extent to which anxiety in the population is related to covid-19 related hospitalizations or deaths. If anxiety can be properly reduced in the population, health care will be safeguarded. Thus, any strategy that reduces anxiety can then be included in evidence-based decision making.

## Introduction

The nocebo effect is any harmful reaction following a negative suggestion. That suggestion can arise from conscious and unconscious meanings that have actions, objects or words. This creates negative expectations that can also harm health (1). When they appear within a medical care context, the Hippocratic Oath “primum non nocere” is compromised.

Anxiety can be seen as a manifestation of a nocebo context. Negative expectations are not in proportion to the real threat. For example, because the anxiety is related to misinterpreting a catastrophe, and mainly driven by metacognitive beliefs (2). For example, people who expected to get the flu before the flu season also had the flu more often afterwards (3).

The psychological stress of the COVID-19 pandemic led some people to experience COVID-19 symptoms, which were not actually related to a COVID-19 infection. This can be seen as a nocebo answer. News and social media may reinforce the negative expectations of larger groups of people by repeating statements that everyone can be affected (4–6). During the 2009-2010 H1N1 influenza pandemic, the tendency to exaggerate the likelihood and severity of infection caused much anxiety that led to a massive psychogenic illness (7).

A fundamental goal during the COVID-19 pandemic is to limit the COVID-19 related demand on healthcare systems and to minimize COVID-19 related deaths. Looking back at another known virus, HIV, it was seen that anxious but healthy people visited the hospital to take an HIV test, often as a reassurance from the anxiety of being infected. In the context of the COVID-19 pandemic, this behavior may inadvertently lead to further spread of hospital acquired (nosocomial) infections and overload the healthcare system. For example, intensive care physicians report an influx of patients who can better contact their GP (8,9). The pressure to test can also lead to higher numbers of false positives. Which further fuels the pandemic anxiety unnecessarily (10).

30 years of research show a clear link between psychological stress and the human immune system (11). It is well understood that chronic stress is a major modulator of immunity and thus directly influences probability of infection (12). For example, colleagues recently found a link between depression and the immune responses measured in the blood after a covid-19 infection (13). It is therefore plausible that populations under stress have a lower resistance, which can be a breeding ground for the SARS-CoV-2 virus (stress as a virus niche).

According to a study in the UK, psychological factors such as stress and an existing mental illness increase the risk of covid-19 hospitalization (14). In the general world population, there is a positive relationship between the psychological well-being for the COVID-19 pandemic and the chance of COVID-19 related survival (15).

This exploratory study aims to determine to what extent the anxiety in the population is related to the number of covid-19 related hospitalizations and deaths.

### Methods

We will collect all studies that assessed anxiety, as measured by a validated scale, during the Covid-19 pandemic in the general population.

A search strategy will take into account the limited resources for this study and will be used to search Pubmed, MedRXiv and PsychRXiv. At PubMed we will use a combination of controlled vocabulary (MeSH terms) and free text words. Three concepts will be combined: “COVID-19 or coronavirus” AND [(validated scales for anxiety or depression) OR (psychological health)] (see Appendix 1 for search strategy).

One researcher (FF) will scan manually the titles and abstracts. Full texts of potentially relevant studies will be collected. Multiple reports for the same study will be linked. FF will screen the full texts for possible inclusion.

Studies will be included if they had at least 100 respondents, used a validated anxiety scale, reported on the general population of a country, and were conducted during the Covid-19 pandemic. For every anxiety measuring scale, the number of studies using it will be counted. Finally, only those studies that reported the most commonly used scale will be included for the analysis.

The data extraction will be double checked by a second researcher (AN). Any non-matching value will be mutually resolved and the resulting value will be adopted after a common agreement.

Because we assume significant inaccuracies in the representativeness of the survey results, we will build an ad hoc imprecision tool with the aim of assigning a weight per study. For this purpose, the final set of included studies will be examined for reported factors significantly impacting the anxiety scores. Their mean absolute impact on the score will be determined. Afterwards, the relative importance per predictor will be determined by calculating the z-score relative to the average impact. We will rank the predictors mentioned in at least half of the studies. Values for these predictors will be extracted.

Official websites of governments will be screened to determine hospitalizations and deaths. In case of missing figures, contact will be made by email. Authors of the included studies will also be asked where relevant data can be found.

Data will be extracted in a spreadsheet. If there were different dates of data collection, the period that is the earliest in the epidemic wave will be chosen. For every study we will calculate the number of new covid-19 related hospitalisations or deaths up to 30 days after the closing data of the studies data collection.

Weighted OLS regression will be used to quantify the relationship of anxiety and hospitalizations / deaths. Weighting consists of (1 / [variance] x [imprecision]) x [population size]). For countries with multiple studies, a one pooled estimate will be calculated by a generic inverse variance meta-analysis. A random effects model will be chosen because we foresee the included studies will have considerable methodological (study design, risk of bias) heterogeneity. A restricted maximum likelihood random effects variance component model will be chosen to allow for residual variance.

Missing study data will be requested from the authors. In case of no response, a second reminder email was sent.

Analyzes will be done with R (16) and data will be imported from Apache OpenOffice spreadsheet (17) with package readODS (18), data manipulation with tidyverse (19), and meta-analyzes with package metafor (20).

## Discussion

This study will test the hypothesis that that population anxiety during the covid-19 epidemic may be associated with covid-19 related hospitalisations or deaths. The results can be used to understand to what extend anxiety in the population can put a stress on the healthcare system. Also to become more critical as to what are possible sources of anxiety. Given the already high societal cost to prevent covid-19 related hospitalisations and deaths, it is crucial to address whether anxiety worsens the situation or not. Latest data from France and Belgium indicate that when the epidemic wave subsides, anxiety also decreases (21,22). The anxiety curve seems to follow the epidemic curve quite well.

### Strength and weaknesses

Our research efforts will be the first to investigate world population anxiety and the relation with convid-19 related burden on public health. We will examine the impact of anxiety across all available evidence. Therefore, we will use an extensive and systematic search for eligible studies. Data extraction will be checked independently by a second researcher. We will cater upon the open science framework. All study processes will be transparent, analysis will be shared, and data will be publicly available.

One possible limitation of the study is that we will use reported study data only, which may not be an accurate representation of actual study conduct: reporting can be poor but study conduct can be good, and vice versa. Secondly, we are evaluating only results from one commonly used anxiety measurement scale, albeit across a comprehensive set of studies. Thirdly, by the nature of the current epidemic, we will likely include many preliminary research findings through preprint, which lessens the robustness of findings. Fourth, This study will be observational. So it does not allow statements about directionality or causality. As studies represent data at the population level, it will not be valid to make statements about the individual chances of people being hospitalized or dying from anxiety. Fifth, precision of findings is likely to be limited as countries register hospitalizations and deaths related to Covid-19 in different ways. Also cross-sectional surveys are rarely representative due to various sources of bias. However, we will use the available data by applying an imprecision weight, calculated according to an ad hoc imprecision tool.

### Implications for research

Following the covid-19 pandemic, governments are trying to take effective measures to protect public health. However, little quantitative evidence is available on the relationship between anxiety and Covid-19 related hospitalizations or deaths. As a result, this psychological phenomenon is largely ignored in policy decisions. However, for decades yet, evidence has shown that anxiety and depression significantly weaken the immune system. The findings of this study will provide insight into the magnitude of the relationship between anxiety and Covid-19 related health harms. By quantifying this relationship, we hope health professionals incorporate these effects into treatment strategies and inform patients of the importance of a calm mind. We also hope that governments incorporate our findings into measures and communications that promote Covid-19 related public health. In addition, the results of this study may encourage a further study that helps clarify the impact of anxiety or other psychological problems on Covid-19 related hospitalization or death.

## Data Availability

data of study results will be published publicly

## Appendix 1

### Validated scales to be searched

**Children’s Anxiety Scale, Depression Anxiety Stress Scale DASS-21, Generalized Anxiety Disorder 7-item GAD-7, Generalized Anxiety Disorder questionnaire-IV GADQ-IV, Geriatric Anxiety Inventory GAI, Hamilton Anxiety Rating Scale HARS, Hamilton Rating Scale For Anxiety HAM-A, Health Anxiety Inventory HAI, health anxiety Whiteley-6, Hospital Anxiety Depression Scale HADS, Overall Anxiety Severity Impairment Scale OASIS, Panic Disorder Severity Scale PDSS, Patient-Reported Outcomes Measurement Information System anxiety scale PROMIS, Severity Measure For Generalized Anxiety Disorder, The State-Trait Anxiety Inventory STAI, Web-Based Depression Anxiety Test WB-DAT**,

### PubMed search strategy

((((“coronavirus”[MeSH Major Topic]) OR (coronavirus[Title/Abstract]) OR (corona virus[Title/Abstract]) OR ((((severe acute respiratory syndrome coronavirus[Title/Abstract]) OR (SARS CoV-2[Title/Abstract])) OR (SARS-CoV[Title/Abstract])) OR (COVID[Title/Abstract]) OR (covid*[Title/Abstract]))))) AND (((Severity Measure For Generalized Anxiety Disorder[Text Word]) OR (Children’s Anxiety Scale[Text Word]) OR (Penn State Worry Questionnaire[Text Word]) OR (Panic Disorder Severity Scale[Text Word]) OR (PDSS[Text Word]) OR ((Health Anxiety Inventory[Text Word]) OR HAI[Text Word]) OR ((Hamilton Rating Scale For Anxiety[Text Word]) OR HAM-A[Text Word]) OR ((Generalized Anxiety Disorder 7-item[Text Word]) OR GAD-7[Text Word]) OR (((Depression Anxiety[Text Word] AND Stress Scale[Text Word])) OR DASS-21[Text Word]) OR ((health anxiety[Text Word]) OR Whiteley-6[Text Word]) OR ((Patient-Reported Outcomes Measurement Information System anxiety scale[Text Word]) OR PROMIS[Text Word]) OR ((Geriatric Anxiety Inventory[Text Word]) OR GAI[Text Word]) OR ((The State-Trait Anxiety Inventory[Text Word]) OR STAI[Text Word]) OR ((Generalized Anxiety Disorder questionnaire-IV[Text Word]) OR GADQ-IV[Text Word]) OR ((Hamilton Anxiety Rating Scale[Text Word]) OR HARS[Text Word]) OR ((Leibowitz Social Anxiety Scale[Text Word]) OR LSAS[Text Word]) OR (((Overall Anxiety Severity[Text Word] AND Impairment Scale[Text Word])) OR OASIS[Text Word]) OR (((Hospital Anxiety[Text Word] AND Depression Scale[Text Word])) OR HADS[Text Word]) OR ((Penn State Worry Questionnaire[Text Word]) OR PSWQ[Text Word]) OR (((Web-Based Depression[Text Word] AND Anxiety Test[Text Word])) OR WB-DAT[Text Word])) OR ((“quarantine”[MeSH Major Topic]) OR ((“social isolation”[MeSH Major Topic]) OR (“loneliness”[MeSH Major Topic])) OR (“psychology”[MeSH Major Topic]) OR (((“mental health”[MeSH Major Topic])) OR (“mental disorders”[MeSH Major Topic])) OR (“anxiety”[MeSH Major Topic]) OR (anxiety[MeSH Major Topic]) OR (depression[MeSH Major Topic]) OR (“stress”[Other Term]) OR (“anger”[MeSH Major Topic]) OR (“grief”[MeSH Major Topic]) OR (“burnout, professional”[MeSH Major Topic]) OR (mental disorder*[Title/Abstract] OR Self-isolation[Title/Abstract] OR social distanc*[Title/Abstract] OR psych*[Title/Abstract] OR mental health[Title/Abstract] OR mental illness*[Title/Abstract] OR stigma[Title/Abstract] OR anxiety*[Title/Abstract] OR anxiety[Title/Abstract] OR anxious[Title/Abstract] OR depression[Title/Abstract] OR depressive[Title/Abstract] OR loneliness[Title/Abstract] OR stress*[Title/Abstract] OR trauma*[Title/Abstract] OR post-traumatic[Title/Abstract] OR posttraumatic[Title/Abstract] OR anger[Title/Abstract] OR mood*[Title/Abstract] OR irritability[Title/Abstract] OR irritable[Title/Abstract] OR emotional disturbance*[Title/Abstract] OR grief[Title/Abstract] OR burned out[Title/Abstract] OR burnout[Title/Abstract])))

#### MedRXiv & PsyArXiv

(isolation OR “mental health” OR “mental illness” OR “mental disorder”) AND (COVID OR covid19) OR (psychology OR psychological OR psychosocial OR anxiety OR depression OR stress or trauma) AND (COVID OR covid19)

## Appendix 2

### Screening Questions for Title and Abstract and Full-text Review Inclusion and Exclusion Criteria

**No: not original human data or a case study or case series**. If it is clear from the title and abstract that the article is not an original report of primary data, but, for example, a letter, editorial, systematic review or meta-analysis, or it is a single case study or case series, then it is excluded. Studies reporting only on animal, cellular, or genetic data are also excluded. Conference abstracts are included.

**No: not a study of any population affected by the COVID-19 outbreak**. If it is clear from the title or abstract that the study is not about any population affected by the COVID-19 outbreak, it is excluded. Studies that include fewer than 100 subjects, are excluded.

**No: not a study which reports mental health symptoms during COVID-19 outbreak**. If it is clear from the title or abstract that the study does not report proportions of participants meeting diagnostic criteria using a validated (anxiety of COVID-19 scale is under validation but will be accepted) diagnostic interview or symptoms (based on a threshold or measured continuously) (e.g., anxiety, depression, stress, loneliness, anxiety, anger, grief, other related) at a delineated event related to COVID-19, then it will be excluded. Delineated events may include pandemic announcements, social isolation regulations, escalation from initial outbreak to peak of outbreak, etc…

**No: not a study which reports mental health symptoms individually for a country**. Aggregate data from multiple countries are not considered. Individual country data must be reported.

**Yes: study eligible for inclusion in full-text review**.

## Appendix 3

### Items to be included in the data extraction

study ID

primary author

email

publication status

DOI

study design

country

population

deaths per 100k

cumulated hospitalizations per 100k

dates of data collection

phase of epidemic

reported weighting procedures

pre-existing mental illness sample %

pre-existing mental illness population %

aged 60 or older sample %

aged 60 or older population %

aged 46-59 sample%

aged 46-59 population %

being female sample %

being female population %

GAD-7 mean N

GAD-7 mean

GAD-7 mean

SD GAD-7 median

GAD-7 median IQR

reported representativeness comments

## Acknowledgements

This research is initiated by FF and has no external funds. Statistical codes and data are - given correct citation of this study - reusable via DOI 10.17605/OSF.IO/GUBQT We will thank study authors for sharing additional information.

## Author information

### Affiliations

#### Independent researcher, Antwerpen, Belgium

Frederik Feys

#### Faculty of Pharmacy, Isra University Jordan/Amman

Abdallah Naser

### Corresponding author

Correspondence to

## Additional information

### Authors’ contributions

FF has the main responsibility of this research and drafted this protocol, developed the search strategy, will search for studies, extract data into Apache Openoffice, carry out the analysis in R, interpret the analysis, and draft the report. AN will double check data extraction forms and will assist in the final write-up of the study manuscript.

## Rights and permissions

This is an article published under the terms of the Creative Commons Attribution License (http://creativecommons.org/licenses/by/2.0), which permits unrestricted use, distribution, and reproduction in any medium, provided the original work is properly cited.

## References

1. Colloca L, Finniss D. Nocebo effects, patient-clinician communication, and therapeutic outcomes. JAMA. 2012 Feb 8;307(6):567–8.

2. Bailey R, Wells A. Is metacognition a causal moderator of the relationship between catastrophic misinterpretation and health anxiety? A prospective study. Behav Res Ther. 2016 Mar;78:43–50.

3. Pagnini F, Cavalera C, Volpato E, Banfi P. Illness expectations predict the development of influenza-like symptoms over the winter season. Complement Ther Med. 2020 May;50:102396.

4. Faasse K, Petrie KJ. The nocebo effect: patient expectations and medication side effects. Postgrad Med J. 2013 Sep;89(1055):540–6.

5. Olagoke AA, Olagoke OO, Hughes AM. Exposure to coronavirus news on mainstream media: The role of risk perceptions and depression. Br J Health Psychol. 2020 May 16;e12427.

6. Farooq A, Laato S, Islam AKMN. Impact of Online Information on Self-Isolation Intention During the COVID-19 Pandemic: Cross-Sectional Study. J Med Internet Res. 2020 May 6;22(5):e19128.

7. Wheaton MG, Abramowitz JS, Berman NC, Fabricant LE, Olatunji BO. Psychological Predictors of Anxiety in Response to the H1N1 (Swine Flu) Pandemic. Cogn Ther Res. 2012 Jun 1;36(3):210–8.

8. Chatterjee SS, Barikar C M, Mukherjee A. Impact of COVID-19 pandemic on pre-existing mental health problems. Asian J Psychiatry. 2020 Apr 18;51:102071.

9. Iacobucci G. Covid-19: Doctors sound alarm over hospital transmissions. BMJ [Internet]. 2020 May 19 [cited 2020 Jul 7];369. Available from: https://www.bmj.com/content/369/bmj.m2013

10. Fenton NE, Neil M, Osman M, McLachlan S. COVID-19 infection and death rates: the need to incorporate causal explanations for the data and avoid bias in testing. J Risk Res. 2020 Apr 24;0(0):1–4.

11. Segerstrom SC, Miller GE. Psychological stress and the human immune system: a meta-analytic study of 30 years of inquiry. Psychol Bull. 2004 Jul;130(4):601–30.

12. Morey JN, Boggero IA, Scott AB, Segerstrom SC. Current Directions in Stress and Human Immune Function. Curr Opin Psychol. 2015 Oct 1;5:13–7.

13. Yuan B, Li W, Liu H, Cai X, Song S, Zhao J, et al. Correlation between immune response and self-reported depression during convalescence from COVID-19. Brain Behav Immun. 2020 May 25;

14. Batty GD, Deary I, Luciano M, Altschul D, Kivimaki M, Gale C. Psychosocial factors and hospitalisations for COVID-19: Prospective cohort study of the general population. medRxiv. 2020 Jun 1;2020.05.29.20100735.

15. Feys F. Is the psychological well-being of a population associated with COVID-19 related survival? medRxiv. 2020 Jun 11;2020.06.05.20123018.

16. R Core Team. R: A language and environment for statistical computing. https://www.R-project.org/. R Foundation for Statistical Computing, Vienna, Austria. [Internet]. 2020. Available from: https://www.R-project.org

17. The Apache Software Foundation. Apache OpenOffice. The Free and Open Productivity Suite [Internet]. Available from: http://www.openoffice.org/

18. Schutten G-J, Chan C, Thomas J. Leeper and other contributors (2018). readODS: Read and Write ODS Files. R package version 1.6.7. [Internet]. Available from: https://CRAN.R-project.org/package=readODS

19. Wickham H, Averick M, Bryan J, Chang W, McGowan LD, François R, et al. Welcome to the Tidyverse. J Open Source Softw. 2019 Nov 21;4(43):1686.

20. Viechtbauer W. Conducting Meta-Analyses in R with the metafor Package. J Stat Softw. 2010 Aug 5;36(1):1–48.

21. Covid-19 : une enquête pour suivre l’évolution des comportements et de la santé mentale pendant l’épidémie [Internet]. [cited 2020 Jul 9]. Available from:/etudes-et-enquetes/covid-19-une-enquête-pour-suivre-l’évolution-des-comportements-et-de-la-sante-mentale-pendant-l-epidemie

22. Demarest. Derde COVID-19 gezondheidsenquête: eerste resultaten [Internet]. sciensano.be. [cited 2020 Jul 9]. Available from: https://www.sciensano.be/en/biblio/derde-covid-19-gezondheidsenquete-eerste-resultaten

